# Does Biological Therapy Protect against Severe COVID-19?

**DOI:** 10.1101/2020.06.21.20136788

**Authors:** Ramon Mazzucchelli, Raquel Almodovar González, Natalia Crespi Villarías, Elena García Zamora, Elia Pérez Fernández, Javier Quirós Donate, Monserrat Pérez Encinas, Patricia Sanmartín Fenollera, María Velasco, Pilar López Serrano, Jose Lázaro Perez Calle, Conrado Fernández Rodríguez, José Luis López-Estebaranz, Pedro Zarco

## Abstract

**Objective:** To estimate COVID-19 infection incidence rate with severe affectation (requiring hospitalization) in patients with biological treatment due to rheumatoid arthritis (RA), psoriatic arthritis (PsA), spondyloarthritis (SpA), psoriasis (Ps), and inflammatory bowel disease (IBD) and compare it with incidence rate in the general population.

**Methods:** Retrospective observational study based on information provided by two administrative databases. One of these two databases contains information on all patients seen in our hospital and diagnosed with COVID-19 infection between March 4th 2020 and April 26th 2020. The other database contains data from patients seen at Rheumatology, Dermatology and Digestive Departments in our hospital who are currently receiving biological therapy. We calculated the crude and age and sex adjusted incidence in both groups. To compare both groups we calculated the Incidence Rate Ratio.

**Results:** There was a total of 2,182 patients with COVID-19 requiring hospitalization. Four patients out of a total of 797 patients receiving biological therapy had contracted COVID-19 and required hospital care. Crude incidence rate of COVID-19 requiring hospital care among the general population was 1.41%, and it was 0.50% among the group receiving biological therapy. Rates adjusted by age and sex in the biological group was 0.45% (CI95% 0.11-4.13). The IRR of the group receiving biological therapy compared to the general population was 0.39 (CI95% 0.14-1, p=0.049).

**Conclusion:** Findings suggest that prior use of biological therapy does not associate with severe manifestations of COVID-19, and it is likely to have a protective effect against them when compared to the general population.

**Key Messages:** *What is already known about this subject?:*

- Covid-19 susceptibility in patients with immune-mediated disorders and receiving treatment with biological therapy is unknown.

*What does this study add?:*

- Severe manifestation incidence rate in patients with immune-mediated disorders receiving biological therapy treatment is not increased when compared to the general population.
- Biological therapies might protect patients from presenting severe COVID-19 manifestations.

*How might this impact on clinical practice?:*

- These data could be used for current recommendations regarding management of patients receiving biological therapies.

**Mini Abstract:** The objective of this study is to analyze the incidence rate of severe COVID-19 requiring hospital care for patients receiving biological therapy and to compare it to the general population. Patients treated with biological therapy have crude and adjusted incidence rates under those of the general population.

**Statement of Human and Animal Rights:** This article does not contain any studies involving human participants or animals that were performed by the authors. For this type of study, formal consent was therefore not required.

## INTRODUCTION

In December 2019, the Chinese government formally announced a severe form of pneumonia that was caused by a new type of coronavirus. It started in Wuhan (Hubei province) and quickly spread all over China, and then all over the world (1). The World Health Organization (WHO) defined this syndrome as Coronavirus Disease-2019 (COVID-19), which was later renamed as severe acute respiratory syndrome coronavirus 2 (SARS-CoV-2) by the ICTV Coronaviridae Study Group (2). SARS-CoV-2 is one of the seven betacoronaviruses and belongs to the coronavirus family (3) that can infect humans and other mammals (4).

Severe pulmonary and systemic inflammatory manifestations observed in COVID-19 led to a hypothesis about a mechanism of hyperinflammation that depends on the host’s response rather than on the direct virus-induced cell damage (5, 6). Certain parallelism with other cytokine storm situations such as macrophage activation syndrome (MAS) or systemic inflammatory syndromes associated with the chimeric antigen receptor T-cell (CAR-T) has been mentioned to support this. This hypothesis has led to the fast introduction of anti-inflammatory and immunomodulatory agents approved for rheumatic disorders in therapeutic strategies against COVID-19. COVID-19 prevalence in patients with previous autoimmune or inflammatory diseases is unknown. Although excess morbidity and mortality has been reported in recent studies (7-9), there seems to not be a higher risk associated to conventional or directed immunosuppression use. It has been hypothesized about a possible preventive or therapeutic effect of certain immunomodulating therapies in these patients. Among them, antimalarials, colchicine, corticosteroids, JAK inhibitors and IL-6/IL-1 receptor antagonist are also being used in special conditions or clinical trials with weak evidence so far (2). However, the risks these drugs carry in the context of viral infections without concomitant antiviral therapies are significant (10-12). It is unknown if immunosuppressive and biological therapies give patients with immune-mediated disease a higher or lower risk of contracting more severe forms of COVID-19, and there is a need for further studies to clarify this (13, 14).

Therefore, the question currently without an answer is whether patients with immune-mediated diseases and treated with biologic agents or immunosuppressants are or are not protected against severe COVID-19 developing.

### Patients and Methods

This is a monocentric cohort observational prospective study and retrospective data collection based on the utilization of two administrative databases. One of the databases contains data on patients diagnosed with COVID-19 seen at *Hospital Universitario Fundación Alcorcon* (clinical diagnosis, and most of them confirmed via PCR test) between March 4^th^, 2020 and April 26^th^, 2020. This period of time covers a timeframe extending before and after the peak of COVID-19 incidence infection in Spain. This database contains data from patients attended at the emergency service whether they were hospitalized or not. Collected data consists of at least data including age, sex, PCR positive or negative, if there was hospitalization, type of discharge (including exitus), admission and discharge dates.

This study has been carried out in Alcorcon, a municipality of the metropolitan area of Madrid (Spain) and a typical dormitory town where roughly half of the total population (169,502 inhabitants at present) works in Madrid. Its population pyramid is typical of an ageing population.

The second database collects data on patients from Rheumatology, Dermatology and Digestive Departments at *Hospital Universitario Fundación Alcorcon* who are currently receiving biologic therapy. Treatments included in this study were as follows: anti-TNF drugs (adalimumab, certolizumab, etanercept, golimumab and infliximab); IL6 inhibitors (sarilumab and tocilizumab); abatacept; anti-IL17 medications (brodalumab, ixekizumab and secukinumab); anti-IL-23 agents (guselkumab and ustekinumab); and others (belimumab, rituximab, tildrakizumab, tofacitinib, baricitinib and vedolizumab). In this database, the variables collected are age, sex, main diagnosis, biological therapies and treatment start date.

Both databases are cross-referenced by medical history number. COVID-19 crude incidence rate adjusted by age and sex was estimated for the group receiving biological therapy and compared to the general population. For this comparison, the biological therapy group’s relative risk (RR) was calculated in relation to the general population. The Municipal Registry of Alcorcon was used to calculate the control group’s (the general population) incidence rates.

Medical records of patients with biological therapy were reviewed to confirm the COVID-19 medical diagnosis. Because PCR test availability was limited, these records only contain data on patients who were attended at *Hospital Universitario Fundacion Alcorcon*, and exclude other cases occurring in the Community of Madrid that were less severe and did not require hospitalization nor were referred to hospital emergency services. Due to a recent study on seroprevalence carried out in Spain (15), it is estimated that COVID-19 prevalence in the Community of Madrid, region where *Hospital Universitario Fundación Alcorcon* belongs, is 11% (https://www.mscbs.gob.es/ciudadanos/ene-covid/home.htm). This means an estimated 18,000 subjects were infected. Nevertheless, only a small part of them (between 10% and 12%) required hospital care. This study assumes that those patients who required hospital care suffered from a more severe illness than those who did not require it.

### Statistical Methods

Data are reported as crude rate adjusted by age and sex and compared to rates of the general population by Incidence Rate Ratio (IRR) with 95% confidence intervals and Chi-squared tests. Mean age in the different groups was compared with nonparametric tests.

Statistical analysis was performed with the statistical packages IBM SPSS 20.0, Epidat 4.2 and Stata 15.

This study was authorized by the Ethics Committee for Clinical Research of the *Hospital Universitario Fundación Alcorcon*.

## RESULTS

There was a total of 2,182 COVID-19 cases requiring hospital care in the Alcorcon municipality. Out of this total, 1,532 cases (70.21%) required hospitalization while the rest only were admitted to the emergency room without hospitalization. 44.9% were women and the mean age was 63.82 years (SD 17.9). There was a total of 265 (12.1%) deaths.

In the area around *Hospital Universitario Fundación Alcorcon*, there were 797 patients who at the time of carrying out this study were receiving treatment with different biological therapies prescribed by the Rheumatology Department (59.7%), Dermatology Department (17.4%) and the Digestive Department (22.8%). Out of this total, 184 patients (23.1%) were diagnosed with rheumatoid arthritis, 173 patients (21.7%) were diagnosed with spondyloarthritis, 150 (18.8%) with Crohn’s disease, 138 (17.31%) with Psoriasis, 105 (13.1%) with psoriatic arthritis, 32 (4.2%) with ulcerative colitis and 15 patients (1.8%) received a different diagnosis. Group therapeutic distribution was as follows: 489 (61.36%) anti-TNF drugs, 87 (10.9%) anti-IL-23 agents, 82 (10.3%) anti-IL-17 agents, 45 (5.6%) anti-IL-6, 21 (2.6%) anti-CD28-CD80-86 and 73 (9.16%) included in the group “Others”. 55.4% were women (compared to 51.7% of the general population, p<0.05) and mean age was 53.78 (SD 15.2) (compared to 43.15 (SD 23.1) in the general population, p<0.05) (Table 1).

**Table 1:** Characteristics, crude rates (%) of COVID-19 in the general population and in patients receiving biologic therapies who required hospital care.

|  | General Population (Denominator) |  |  | n' | Cases Covid-19 (Numerator) |  |  | COVID-19 rate (%) | IRR (CI95%) vs Reference |
| --- | --- | --- | --- | --- | --- | --- | --- | --- | --- |
|  | n (%) | Mean age (SD) | Sex (Female%) |  | Mean age (SD) | Sex (Female%) | Exitus (%) |  |  |
| General population (ref) | 169.502 | 43.15 (23.1) (ref) | 51.7 (ref) | 2,178 | 63.82 (17.9) | 44.9 | 265 (12.1) | 1.28 | 1 (reference) |
| Biologic therapy | 797 | 53.78 (15.2)* | 55.45* | 4 | 55 (17.45) | 25.0 | 1 (25%) | 0.5 | 0.39 (0.14-1)* |
| **By diagnosis** |  |  |  |  |  |  |  |  |  |
| rheumatoid arthritis | 184 (23.1) | 63.29 (14.6)* | 77.71* | 1 | 44.46 (-) | 0.0 |  | 0.54 | 0.42 (0.06-3) |
| psoriatic arthritis | 105 (13.17) | 54.22 (14.01)* | 61.9* | 1 | 46.5 (-) | 100.0 |  | 0.95 | 0.74 (0.10-5.30) |
| spondyloarthritis | 173 (21.7) | 51.46 (12.4)* | 50.28 | 0 |  |  |  | 0.00 |  |
| psoriasis | 138 (17.3) | 53.64 (13.9)* | 38.4 | 2 | 65.48 (22.1) | 0.0 |  | 1.45 | 1.12 (0.28-4.57) |
| Chron's disease | 150 (18.8) | 45.50 (15.4)* | 44.66 | 0 |  |  |  | 0.00 |  |
| ulcerative colitis | 32 (4.1) | 48.28 (11.1)* | 46.87 | 0 |  |  |  | 0.00 |  |
| other | 15 (81.8) | 56.63 (13.8)* | 80* | 0 |  |  |  | 0.00 |  |
| **By department** |  |  |  |  |  |  |  |  |  |
| Rheumatology | 476 (59.7) | 56.83(14.6)* | 64.49* | 2 | 45.48 (1.44) |  |  | 0.42 | 0.32 (0.08-1.30) |
| Dermatology | 139 (17.4) | 53.55(13.9)* | 38.12* | 2 | 65.48 (22.1) |  |  | 1.44 | 1.12 (0.28-4.53) |
| Digestive | 182 (22.8) | 45.99 (14.8)* | 45.05 | 0 |  |  |  | 0.00 |  |
| **By therapy** |  |  |  |  |  |  |  |  |  |
| anti-TNF | 489 (61.3) | 52.66 (15.5)* | 52.53 | 2 | 62.78 (25.9) | 0,0 |  | 0.41 | 0.32 (0.08-1.27) |
| anti-IL23 | 87 (10.9) | 51.33 (13.3)* | 43.68 | 1 | 49.8 (-) | 0,0 |  | 1.15 | 0.89 (0.12-6.49) |
| anti-IL17 | 82 (10.3) | 52.9 (13.8)* | 53.66 | 1 | 46.5 (-) | 100,0 |  | 1.22 | 0.95 (0.13-6.82) |
| anti-IL6 | 45 (5.65) | 63.28 (12.2)* | 91.11* | 0 |  |  |  | 0.00 |  |
| anti-CD28-CD80-86 | 21 (2.63) | 68.10 (15.7)* | 80.95* | 0 |  |  |  | 0.00 |  |
| Others | 73 (9.16) | 55.15 (13.9)* | 61.64 | 0 |  |  |  | 0.00 |  |

\*p<0.05. (\*) Incidence Rate Ratio (IRR), confidence intervals, and all comparisons were between each group and the population of reference

Out of the total of 797 patients receiving biological treatment, only four of them (0.18%), three men and one woman, had COVID-19 requiring hospital care. Two patients were in treatment with etanercept, one patient with guselkinumab and one patient with ixekizumab. Main clinical characteristics of these patients are shown in Table 2.

**Table 2:** Characteristics of patients requiring hospital care while receiving biologic therapy

| **Patient** | **1** | **2** | **3** | **4** |
| --- | --- | --- | --- | --- |
| **gender** | Male | Male | Female | Male |
| **age (years)** | 44 | 49 | 46 | 81 |
| **stay (days)** | 11 | 46 | 2 | 15 |
| **Antecedents** |  |  |  |  |
| HTA | no | no | no | no |
| obesity | no | yes | no | no |
| tobacco | no | yes | no | yes |
| venous thrombosis | yes | no | no | no |
| cerebral palsy | yes | yes | no | no |
| **Diagnosis (reason for biologic therapy)** | RA | Psoriasis | PsA | Psoriasis |
| **Biologic therapy** | etanercept | guselkinumab | ixekizumab | etanercept |
| **Concomitant treatment** |  |  |  |  |
| methotrexate | yes | no | yes | no |
| hydroxychloroquine | no | no | no | no |
| corticosteroids | no | no | no | no |
| **CCU admission required** | no | yes | no | no |
| **Exitus** | no | no | no | yes |

COVID-19 crude incidence rate requiring hospital care in the general population was 1.28%, while in the biological therapy cohort it was 0.5%. Figure 1 shows crude rates by age ranges. Rates were adjusted by age (16) and sex: 1.42% (CI95% 1.36-1.48) in the general population and 0.46% (CI95% 0.12-4.13) for the biological therapy group. IRR between the biological therapy cohort and the general population was 0.39 (CI95% 0.14-1; p<0.05).

**Figure 1:**
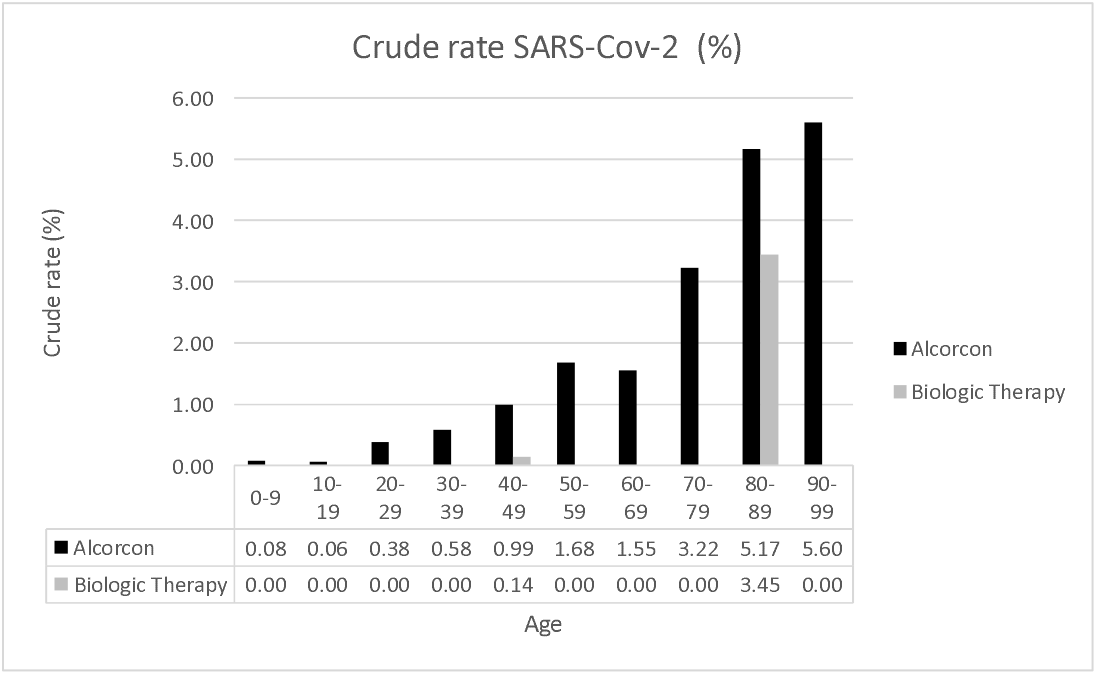
Crude rates by age ranges in Alcorcon: General population and patients with biological therapy.

## DISCUSSION

Using a systematic approach, this study analyzes severe COVID-19 incidence (requiring hospitalization) comparing the cohort of patients treated with biological therapies in Rheumatology, Dermatology and Digestive Departments at *Hospital Universitario Fundacion Alcorcon* with the general population.

The main finding is a lower incidence of severe COVID-19 in the cohort of patients with biological therapy than in the general population (IRR 0.39 (CI95% 0.14-1; p<0.05). Furthermore, this finding is reinforced by the fact that the mean age in the cohort with biological therapy is 10 years older than the mean age in the general population, and it is known that age is the most important risk factor for developing a severe form of the disease (16, 17). On the other hand, there is a higher proportion of women in the biologic therapy cohort (55.4% vs 51.7%; p<0.05)(16, 17). The low number of patients with severe COVID-19 in the biological therapy cohort (only four patients) does not allow analysis of other related aspects like, for example, to analyze which factors are associated with a severe form of the disease. However, it is worth noting that 2 out of the 4 patients had a severe neurological disorder (cerebral palsy) which is also a known risk factor (16).

At present, it is still unclear if patients with immunosuppressive therapy are more likely to get COVID-19 than the general population, and in the case that these patients get it, if these biological therapies lead to a higher rate of complications, such as secondary bacterial pneumonia or acute respiratory distress syndrome (ARDS). Nevertheless, limited data available at the moment suggest that this is not the case, because the host’s innate immune system seems to be the main driver of pulmonary inflammation (1, 6). The retrospective Monti *et al*. (18) study based on a survey given to 13 patients with RA or AS and receiving biological or synthetic therapy who were positive or had highly suggestive characteristics or had known contact with a COVID-19 infected subject, reported no cases of severe respiratory complications or death, and only a 65-year old patient required hospitalization to receive supplemental oxygen (low flow) (18). Furthermore, in the Monti *et al*. study, none of the 700 patients hospitalized due to severe COVID-19 were receiving targeted biological or synthetic therapy, which suggests that patients with immunomodulating therapy may not be at a higher risk or their lives more in danger in comparison to the general population (18). A recent audit on critical care units in UK (https://www.icnarc.org/) showed that out of the 775 patients admitted with symptoms related to COVID-19, only 3% (22 patients) were considered as immunocompromised before admission in comparison to 8.8% of patients admitted due to viral pneumonia before this ongoing COVID-19 pandemic (19). Data published in China do not report on any rheumatologic diseases or immunosuppressive therapy as an important risk factor for severe COVID-19 (17). A recent publication by Haberman *et al*. reported 86 patients with immune-mediated inflammatory disease (including RA, AS, PsA, Ps and IBD) with confirmed or highly suspected COVID-19 infection (20). Hospitalization incidence in that cohort was 16%. This group was larger than the non-hospitalized cohort and had a more elevated incidence of comorbidities such as chronic obstructive pulmonary disease and diabetes (20). Surprisingly, a lower percentage of the hospitalized group received biological therapies or JAK inhibitors in comparison with the non-hospitalized group, while oral glucocorticoids, hydroxychloroquine and methotrexate use was greater (20). Another study, carried out in Spain by Michelena *et al*. analyzes COVID-19 incidence in a cohort of 959 rheumatic patients, children and adults, receiving biological therapy or JAK inhibitors (21). They only found 11 cases with PCR confirmation, all of them adults, six of them with pneumonia (21). When comparing this cohort incidence rate with the general population, they obtained similar results (0.48% vs 0.58% in the general population) (21).

There are also recent studies reporting excess morbidity and mortality in patients with immune-mediated diseases (7-9). In another study carried out in Spain, De Pablos *et al*. observed that certain inflammatory arthritis, polymyalgia rheumatica, giant cell arteritis and autoimmune diseases (not SLE) demonstrate an increased risk for more severe COVID-19 manifestations (9). Furthermore, the De Pablos *et al*. study reported patients with inflammatory arthritis (not RA) treated with biological therapies and JAK inhibitors having more risk (OR 1.6 CI95% 1.23-2.1) than the general population for severe manifestation (9). It is worth noting that the De Pablos *et al*. study results are not adjusted by age and sex and probably differences observed are due to the age factor, rather than to diseases and treatments under consideration.

The development of a database, like the COVID-19 Global Rheumatology Alliance, should increase knowledge over the next months (22). So far, this registry has allowed for publication of reviewed data from 110 patients with rheumatic disease and diagnosed with COVID-19 which reports about their rheumatic diagnosis, drugs, COVID-19 symptoms and comorbidities. Although 35% of these patients were hospitalized and 5% died, it is again not possible to extrapolate if disease severity is related to rheumatologic diagnosis/drugs or other comorbidities on the basis of these early data (13).

As for this study, there are limitations. Only cases attended in the emergency service and requiring hospitalization have been identified, although there is knowledge of an unknown, but significant, number of rheumatic patients with mild COVID-19 whose detection was based on self-reports and phone consultations, but without PCR test confirmation. The real incidence of severe and non-severe cases among rheumatic patients, but also for the non-rheumatic population of reference, could only be reliably estimated with seroprevalence studies. One of the main limitations of this study is the low number of COVID-19 cases in the biological therapy cohort, which conditions the ability to analyze factors associated to developing a severe form of the disease.

Taking all these limitations into consideration, definite conclusions cannot be drawn due to the limited number of cases. However, this study’s results suggest that patients with immune-mediated diseases and receiving biological therapy do not show higher risk for developing severe manifestations of COVID-19, and according to our data, it is very likely that this risk is lower than for the general population. In any case, it is necessary to confirm these results with a larger case series including a greater number of patients receiving biological therapy.

## Data Availability

The availability of the database will be on request to the

## Author contributions

RM, PZ and NCV designed research; EPF, RM and NCV analysed data. MPE, JLPC, PLS, EGZ, PSF, MV, CFR, JLE and PZ contributed with the database of their respective units; RME, JQD, RAG and NCV wrote the paper.

## Acknowledgements

The authors thank the staff of the Research Unit of the Spanish Society of Rheumatology for their support in the editing and translation of the manuscript. To my good friend Caligula who faithfully accompanies me in my research work.

